# Designing a Clinical Decision Support Tool to Promote Timely Palliative Care in Heart Failure

**DOI:** 10.64898/2026.01.20.26344491

**Authors:** Yutong Men, Nathan Wright, Giselle O’Connor, Hargun Kaur, Ella Suh, Jenny Chen, Bashar Kadhim, Tariq Ahmad, Michael Beasley, Nora Segar, Jonathan Quang, Mona Sharifi, Shelli Feder

**Author notes:** Corresponding Author: Yutong Men, HBSc, Yale School of Public Health, 60 College Street, New Haven, CT 06510, USA. Co-corresponding authors: Shelli Feder, PhD, FNP-C, ACHPN, FPCN, FAHA, FAAN, Yale School of Nursing, 400 West Campus Drive, Orange, CT 06477, USA.

## Abstract

**Background:** Palliative care improves quality of life and reduces healthcare utilization for people with heart failure, yet referrals remain inconsistent and delayed. Clinical decision support (CDS) offers a promising strategy to facilitate timely palliative care, but no CDS tool currently exists to specifically support palliative care decision-making in this population.

**Methods:** Guided by the User-Centered Framework for Implementation of Technology (UFIT), we conducted a qualitative descriptive study of focus groups and interviews with referring (i.e., hospitalists, cardiologists) and palliative care clinicians across two hospitals in an academic health system. Using rapid qualitative and content analysis, we identified needs, contextual factors, and design requirements to inform CDS tool development for timely palliative care.

**Results:** Clinicians (n=25) identified pain points in the current workflow, clinicians’ goals and expectations for earlier referral, and barriers to timely palliative care, such as timing uncertainty. Informational needs included prognostic and clinical data. Context reflected clinicians’ prior experiences with CDS tools, where they reported few options specific to palliative care and disliked interruptive “pop-up” alerts not tailored to clinical contexts or workflow. CDS requirements included incorporation of objective markers of clinical deterioration, tailored recommendations and workflow integration based on clinical acuity and palliative needs, and a clear, visually appealing design.

**Conclusions:** Clinicians identified critical information for a CDS tool to promote timely palliative care for patients with heart failure. These findings directly inform the design, workflow, implementation strategies, and future pilot testing of our palliative care CDS tool and subsequent clinical trial.

**Clinical Perspective:** *What is new?:*

- This study is the first to apply the UFIT framework to guide the development of a CDS tool to promote timely palliative care among patients with heart failure.
- This work advances palliative care innovation by showing how a CDS tool can address barriers through integration of clinical and contextual insights.

*What are the clinical implications?:*

- This qualitative study informs the development of a clinical decision support tool to promote guideline-concordant palliative care in heart failure.
- Findings underscore that successful implementation of CDS tools depends on the tool’s alignment with established clinical workflows to enhance usability, support clinician adoption, and minimize workflow disruption.

## INTRODUCTION

Heart failure is a progressive, life-limiting condition, projected to impact over eight million people in the United States by 2030.^1^ As the disease advances, hospitalizations and readmissions become increasingly common, where up to 80% of patients are hospitalized in the final six months of life.^2^ These hospital-based episodes account for the majority of clinical care costs in heart failure, which are expected to approach $70 billion by 2030.^1^ Severe and persistent symptoms are common among this population, with levels of distress comparable to those experienced by patients with advanced cancer.^3^

Palliative care is an essential component of evidence-based, guideline-directed care for people with heart failure.^1, 4^ Patients with heart failure who receive palliative care experience lower healthcare utilization near the end of life, including fewer emergency department visits, intensive care unit (ICU) stays, and hospitalizations.^5^ They are also more likely to engage in advance care planning and to receive timely referrals to hospice.^6–9^ However, although 72% of hospitals in the United States report having palliative care programs,^10^ fewer than 20% of individuals with heart failure ultimately receive these services.^11^ Unclear criteria for when to refer, variability in provider awareness and attitudes towards palliative care, and the relative lack of evidence-based interventions to promote timely palliative care referral contribute to its poor usage among this population.^12–15^

Clinical Decision Support (CDS) tools offer a promising approach to bridging this knowledge-to-practice gap, improving adherence to clinical practice.^16^ While CDS tools to support palliative care delivery are emerging, many are narrowly focused on populations with cancer or have demonstrated limited acceptability and feasibility.^17^ Moreover, the successful adoption of CDS tools in general has been historically challenging due to issues with workflow disruption, alert fatigue, clinician resistance, and provider burnout. Critically, no CDS tool currently exists to specifically support timely palliative care decision-making specifically for people with heart failure.

To address this gap, we aimed to develop and pilot test an evidence-based CDS tool to increase palliative care among people with heart failure, beginning within inpatient settings. We present findings from the qualitative phase of the project, seeking to inform CDS development by identifying key needs, contextual factors, and design requirements from the perspective of end users and palliative care, ensuring that feedback directly informed the tool’s design to enhance usability, efficiency, and satisfaction.

## METHODS

### Overview and Conceptual Framework

This study and its reporting adhered to the Standards for Reporting Qualitative Research.^18^ The overall design of our larger pilot study is guided by the User-centered Framework for Implementation of Technology (UFIT), a theoretically grounded pragmatic framework that addresses barriers to CDS implementation by jointly attending to technology design and implementation within existing clinical workflows to enhance usability and adoption.^19^ UFIT incorporates multiple models to ensure stakeholder engagement throughout the design process, aligns CDS functionality with real-world sociotechnical contexts, and translates identified barriers into actionable design refinements and tailored implementation strategies, supporting future dissemination and sustainability.^19^ UFIT integrates complementary frameworks, which include the User-Centered Design, Human Factors/Ergonomics, and the updated Consolidated Framework for Implementation Research.^19^

This paper focuses on the first three steps of UFIT: defining the need, context, and requirement for the CDS. These early steps are critical to informing the subsequent phases of design and testing. By grounding this formative work in UFIT, we hope to ensure that the resulting CDS tool and implementation strategies are well aligned with clinical realities and positioned for successful uptake.

### Study Setting and Participants

We conducted a qualitative descriptive study comprised of focus groups and interviews with clinicians likely to encounter or be affected by the CDS tool. Our sampling frame included clinicians who could potentially refer patients with heart failure to palliative care (i.e., hospitalists and cardiologists) and palliative care clinicians who may receive these referrals at two urban hospitals within a large academic health system in the northeastern United States. These hospitals have inpatient capacities of 1,030 and 511 beds, respectively.

Purposive and chain-referral sampling strategies were used to obtain a diverse range of stakeholder perspectives across the two hospital sites.^20^ Clinicians were identified by hospital leaders and administrators, who assisted with outreach. Eligible clinicians received an email invitation that included a brief description of the study and a link to a REDCap survey, which included study background information, consent documentation, a brief demographic questionnaire, and a link to schedule their session. Referring clinicians received a $75 gift card for participating in a focus group. Palliative care clinicians received a $50 gift card for completing an interview.

Inclusion criteria for referring clinicians included having ordering privileges and/or being a member of an admitting team caring for hospitalized patients with heart failure. For palliative care clinicians, eligibility required being a member of the palliative care team involved in the delivery of inpatient palliative care. This study was approved by Yale University’s Institutional Review Board (IRB# 2000038269).

### Focus Group/Interview Guide Development

Our semi-structured focus group and interview guides were informed by the UFIT framework, and were designed to: 1) identify the cognitive and affective needs of clinicians related to the CDS, 2) understand the context of the CDS by exploring the facilitators and barriers to decision-making around referral to palliative care within the broader healthcare system, and 3) define the requirements of the CDS, and identify what changes are needed to improve care. For palliative care clinicians, we also included questions about potential unintended consequences and how the CDS might impact their workflows.

### Data collection

A total of 25 clinicians participated in 5 focus groups for referring clinicians or interviews conducted between January and March 2025. Verbal informed consent was obtained from all participants. All sessions were held remotely via Zoom by a qualitative researcher and trained research team members. We conducted focus groups, rather than individual interviews, with most referring clinicians to account for our compressed timeline and enhance efficiency. Focus groups also enabled us to gather a broader range of input from these key end users of the CDS tool. The group setting allowed clinicians to build on each other’s ideas, facilitating a more generalizable understanding of referral practices and generating richer insights to inform CDS design. Focus groups and individual interviews lasted approximately 60 minutes and 45 minutes, respectively. Data collection and analysis occurred concurrently; we stopped interviewing additional participants when successive interviews yielded no new insights and data saturation was achieved.

### Data Analysis

All focus group and interview sessions were audio-recorded, transcribed verbatim, and de-identified. We chose a Rapid Qualitative Analysis (RQA) approach – a cost-effective and time-efficient approach that produces results consistent and comparable with those of traditional qualitative methods ^21^ – due to the condensed timeline of the larger study and the need to quickly generate findings to inform the design of the CDS tool. We developed an RQA template a priori, based on the UFIT conceptual framework, interview questions, and the specific informational needs related to CDS development. Each transcript was independently analyzed by two members of the research team using the RQA template (with designated sections for analyzing Need, Context, and Requirement). The resulting summaries were then merged to ensure comprehensive data capture. Any discrepancies were discussed and resolved during regular team meetings. Once finalized, all merged summaries were compiled into a matrix spreadsheet organized by focus group or interview ID and focus group/interview topics.

We then conducted a directed content analysis of the aggregated summaries.^22^ This approach allowed us to systematically examine and synthesize the data within the UFIT framework steps of Need, Context, and Requirement. Two researchers performed the analysis by reviewing the merged summaries within each domain and examining how perspectives converged or diverged across interviews and focus groups. The resulting synthesis was subsequently reviewed and refined in collaboration with the broader research team to enhance rigor and reduce bias. Workflows, barriers, and facilitators, as well as proposed intervention requirements, were synthesized into a process map (Figure 1). The directed content analysis allowed us to move beyond descriptive summaries generated through RQA toward higher-level findings that directly informed the organization of the Results section and the design of the CDS tool. Author Y.M. had full access to all the data in the study and takes responsibility for the integrity of the data and the accuracy of the data analysis.

**Figure 1.**
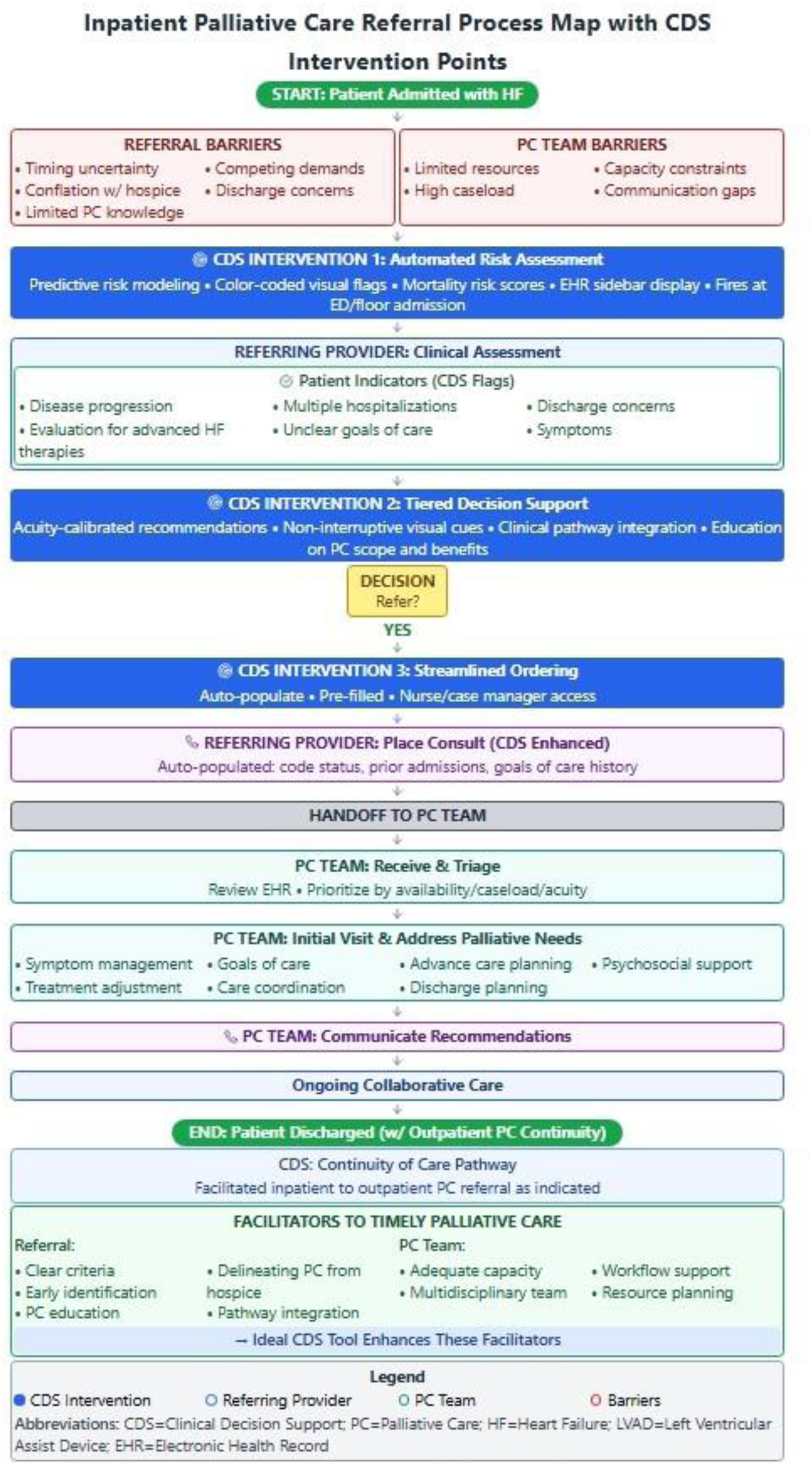
Process map of inpatient palliative care referral for heart failure patients. Blue boxes indicate CDS intervention points addressing barriers for timely PC.

## RESULTS

In total, 18 referring clinicians participated in focus groups and 7 palliative care clinicians in interviews (Table 1). The 25 participating clinicians (mean age 36.9 years, 60% female) were predominantly White (52%) or Asian (32%) and included physicians (attendings, fellows, residents), advanced practice providers, and a social worker.

**Table 1.**
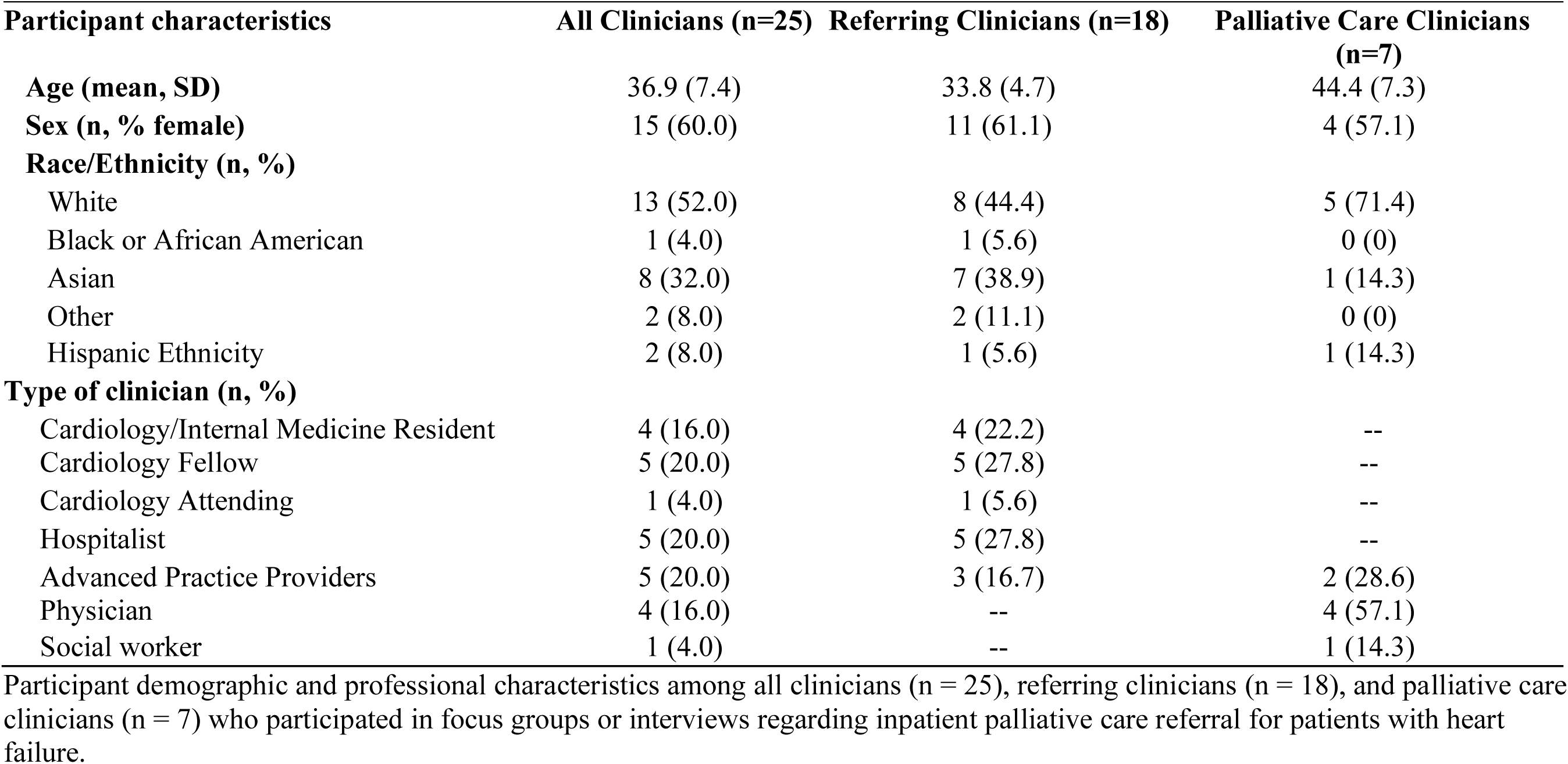
Participant Characteristics by Clinician Role.

Our findings focus on three key areas that inform the design of a CDS tool to support timely palliative care for patients with heart failure: 1) Need – the current workflow for palliative care referral, clinicians’ goals and expectations for referring people hospitalized with heart failure to palliative care, and the perceived barriers and facilitators, as well as the informational needs for palliative care referral; 2) Context – clinicians’ prior experiences with CDS tools; and 3) Requirements – the clinical criteria required for palliative care referral, preferred features and functionality of the CDS tool, strategies for integrating the tool into existing workflows, and identification of the most appropriate end users and timing to encounter the tool.

### Needs

#### Current workflow (process of referral)

Clinicians described a multi-step process by which people hospitalized with heart failure were referred to palliative care during hospitalization (Figure 1). Clinicians noted that referrals were typically prompted by clinical markers of disease progression and health system utilization but were often impeded by barriers such as uncertainty about when to refer; limited knowledge of palliative care and its scope; miscommunication between primary and palliative care teams, patients, and families; and concerns over palliative care team workload.

Referring clinicians placed a consult order in the electronic health record (EHR), which was received and triaged by the palliative care team based on availability and caseload. Additional phone or text communication was often needed to clarify expectations, coordinate timing, and tailor involvement to patient needs. Following the consultation, palliative care clinicians typically reengaged the referring team to share recommendations, such as symptom management, family meetings, or treatment adjustments.

#### Goals and expectations

Although clinicians strongly emphasized the importance of early palliative care for patients with heart failure, in practice, this was not often executed. Clinicians reported that palliative care was consulted primarily when patients were critically ill, rather than earlier in the disease course when longitudinal support could have been more impactful. Uncertainty about the ideal timing to initiate palliative care was frequently mentioned. Clinicians often described conflating palliative care with hospice, a misconception that was a major barrier to timely involvement (Figure 1).

> *“On average, we refer to Pal[liative] Care a little too late. And we involve them when they’re already looking at hospice.”(Referring Clinician)*

Referring clinicians were often unsure of the benefits or function of palliative care, which delayed referral initiation and, in some cases, prolonged hospital discharge. Clearer definitions of palliative care’s role and referral criteria were viewed as essential for promoting timely engagement. Delays were also linked to practical constraints, including limited time for goals-of-care discussions, competing clinical demands, and worries that a palliative care consult might slow discharge planning. Some clinicians also hesitated to introduce palliative care late in a hospitalization out of concern that it could confuse families or disrupt existing care plans, leading to missed opportunities for timely referral.

#### Informational needs for referral to palliative care

Access to relevant clinical information was described as essential when initiating and responding to an inpatient palliative care referral (Table 2). For referring clinicians, such information played a central role in identifying appropriate candidates for palliative care involvement. Commonly cited referral facilitators focused on life expectancy, healthcare utilization (i.e., frequency of hospitalizations/readmissions, length of stay), and eligibility for advanced therapies (i.e., Left Ventricular Assist Device [LVAD] or transplant) (Figure 1). Clinicians reported using these factors to assess disease trajectory and treatment complexity, which guided their decision-making about whether palliative care would be beneficial.

**Table 2.**
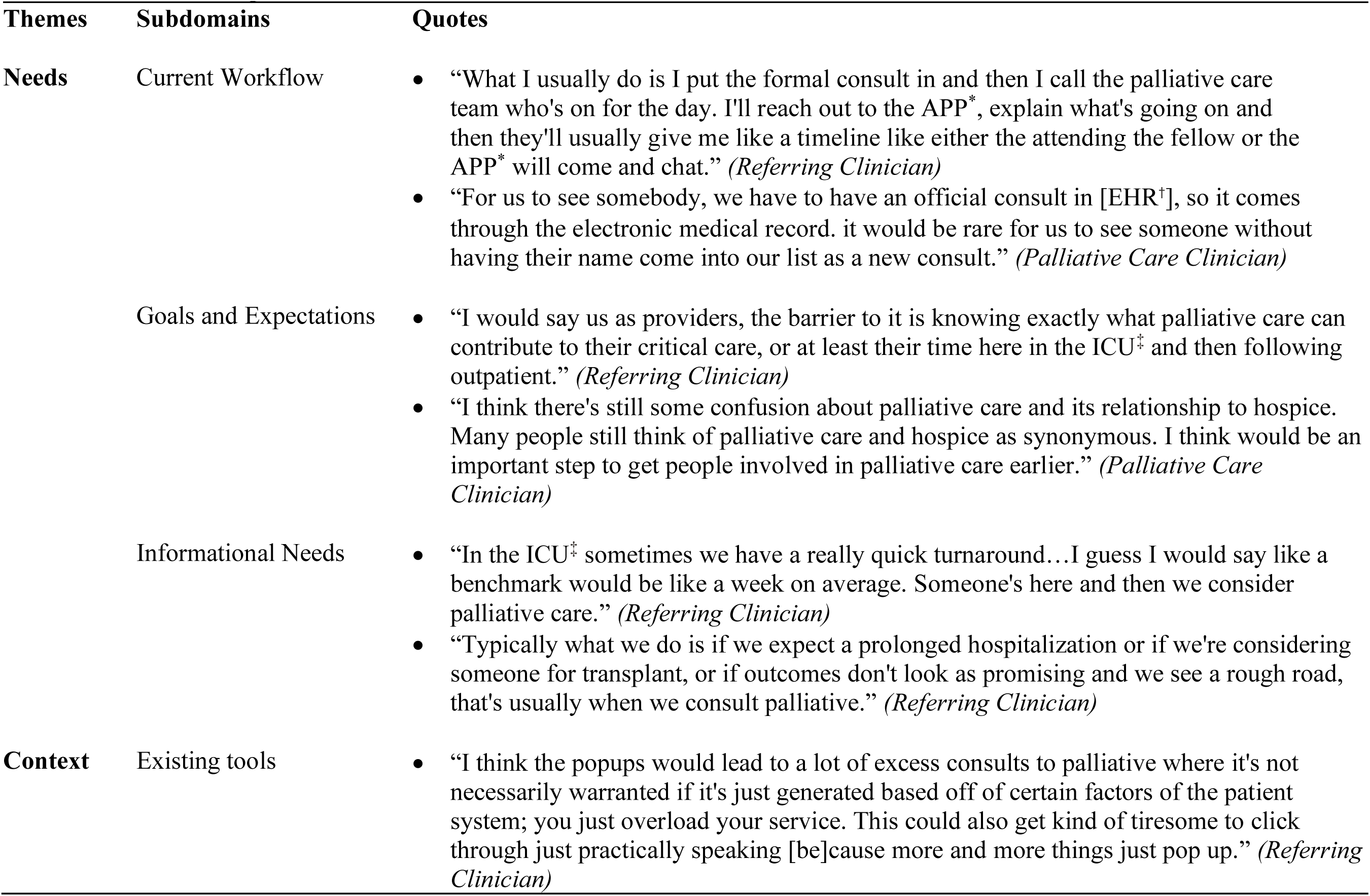
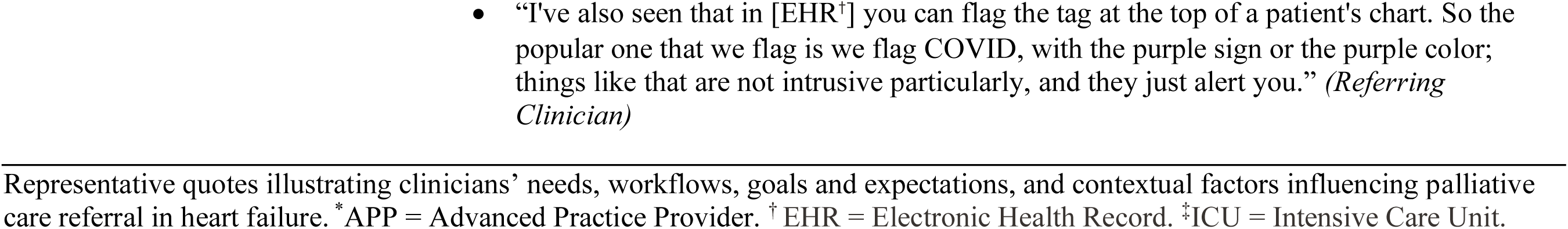
Clinician Perspectives on Needs and Context.

### Context

#### Existing tools

When asked about their experiences with existing CDS tools in the current system and workflow, clinicians most often described clinical pathways. These pathways provided recommended management steps based on a patient’s admission diagnosis or problem list and incorporate structured order sets, which in some cases incorporate order sets that prompt palliative care.

Clinicians expressed conflicted experiences with specific design features of existing CDS tools (Table 2). While some clinicians recognized CDS as helpful, most clinicians expressed negative attitudes toward interruptive alerts that “pop up” automatically within workflows. These were described as burdensome, contributing to alert fatigue, and leading to frequent dismissal of potentially important prompts. Clinicians emphasized that excessive alerts disrupted workflow and risked desensitizing users to critical information, particularly given the already information-dense nature of the EHR interface.

> *“I would caution against what we are increasingly seeing among physicians, as we call alert fatigue. Literally every time I’m opening a patient chart, there’s an alert.” (Referring Clinician)*

Clinicians raised concerns that implementing automated interruptive alerts with referral prompts could inadvertently drive excessive palliative care consults, potentially overwhelming the already resource-limited palliative care service (Table 2). Clinicians also described experiences with color-coded flags within the EHR (e.g., the purple COVID-19 status flag), which appear prominently in the patient chart without requiring user interaction. Clinicians noted that such visual indicators effectively convey key clinical information while being less intrusive and integrate well into their workflow.

#### Requirements

##### Patient Characteristics

Clinicians described a range of patient characteristics they consider when deciding whether to refer patients with heart failure to palliative care (Table 3). Two key features were repeatedly mentioned: mortality risk and healthcare utilization. Clinicians emphasized the need to identify patients who were not imminently dying but remained at significant risk. Healthcare utilization was another major factor influencing decision-making. Clinicians noted that frequent or repeated hospitalizations, particularly for the same clinical issue, often signaled an unmet need for palliative care. Clinicians also highlighted that repeated admissions are associated with higher mortality risk and that early palliative involvement could help reduce readmissions, improve patient education, and enhance care coordination.

> *“Every readmission increases mortality risk by a certain percentage. If I see a patient readmitted twice or more than that, I think it would be a good idea to consult palliative care.” (Referring Clinician)*

**Table 3.**
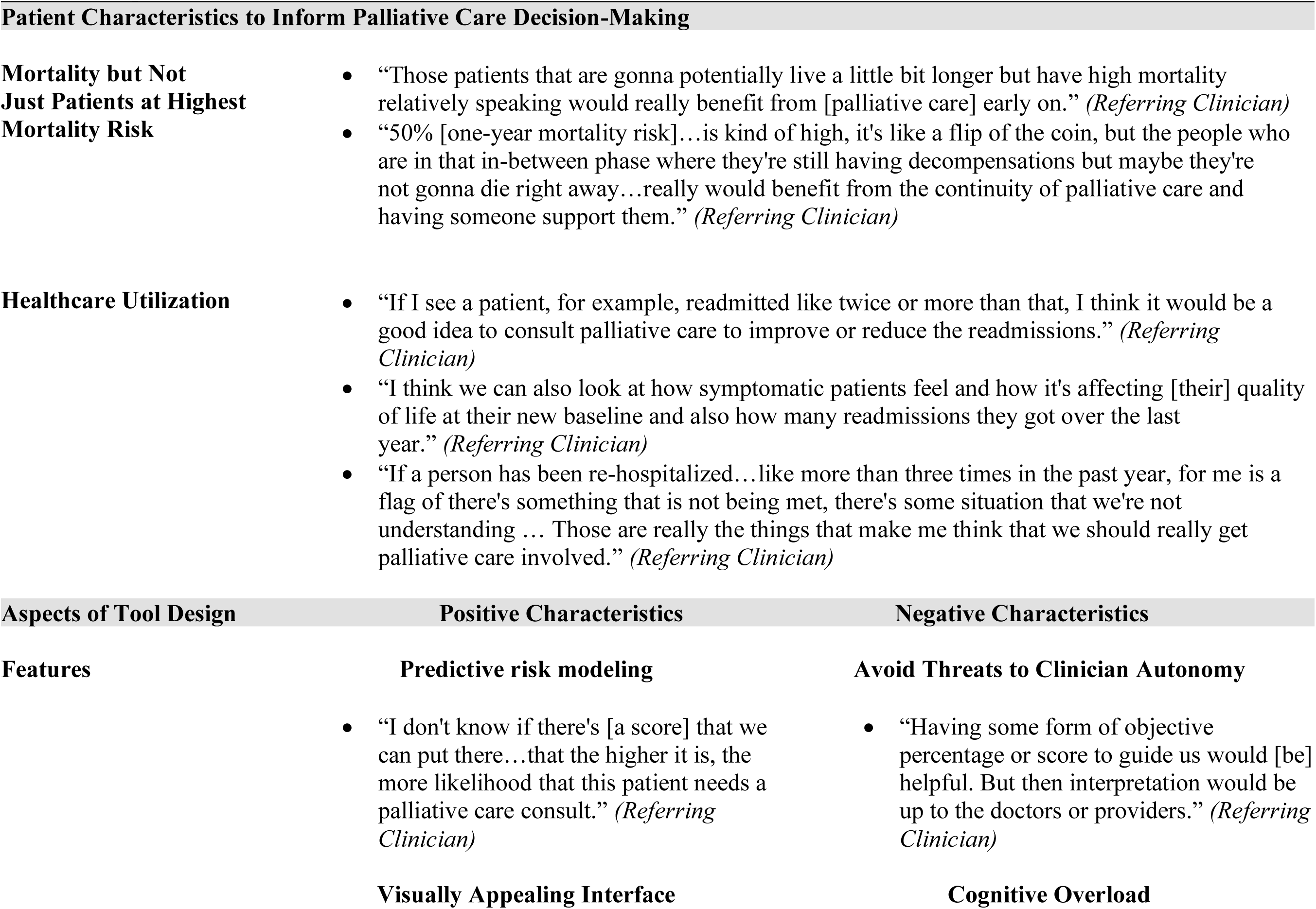
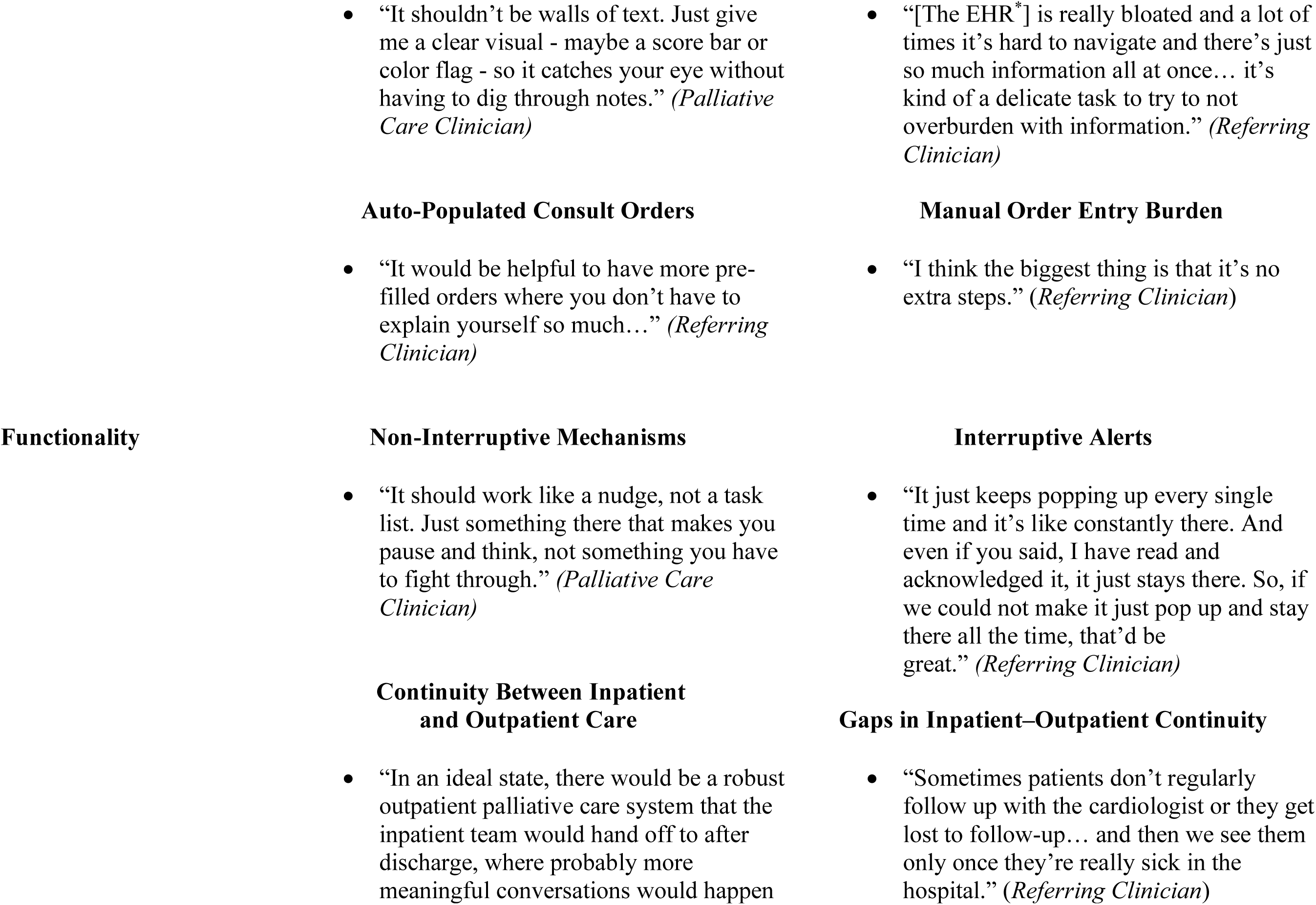
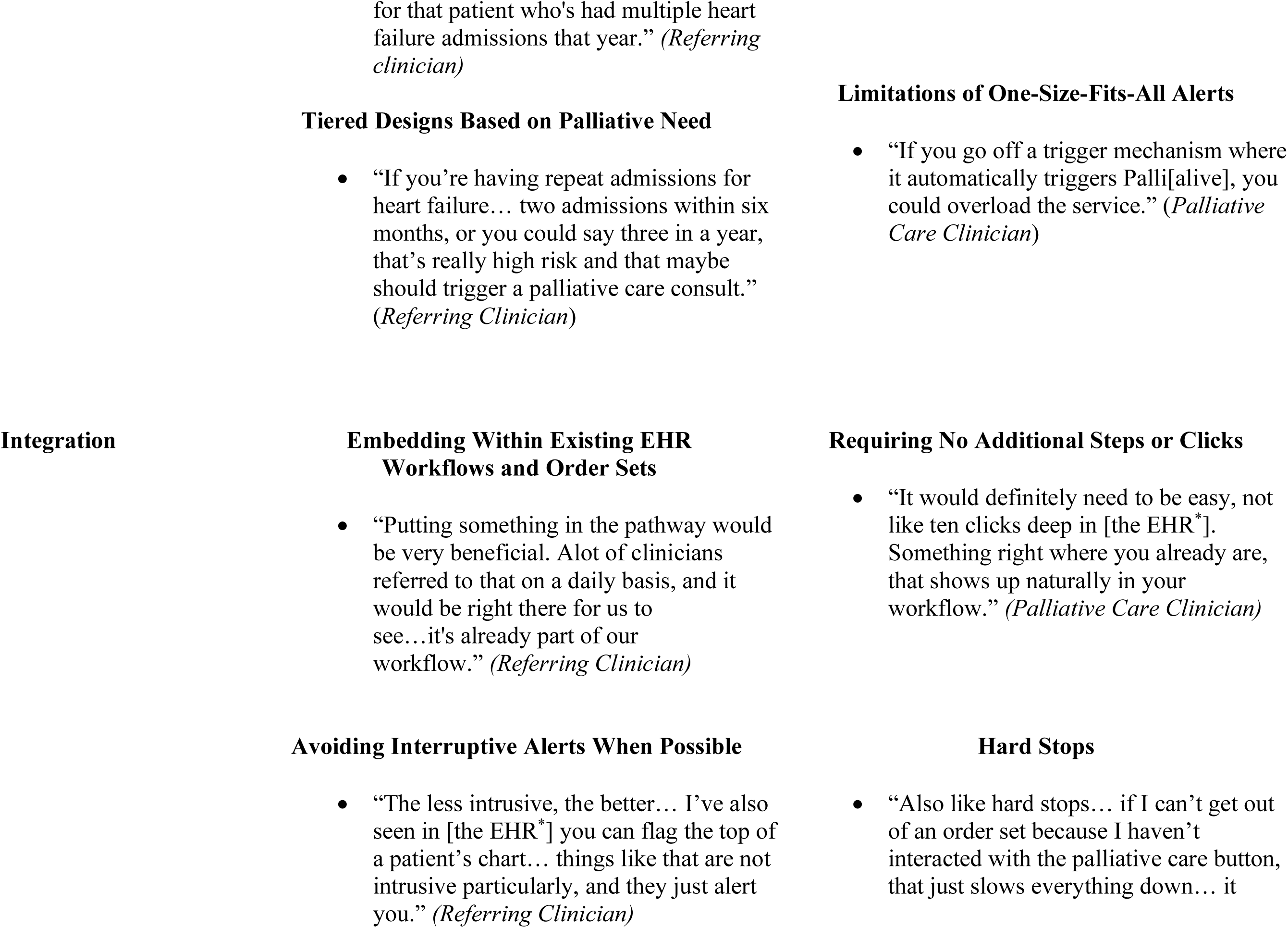
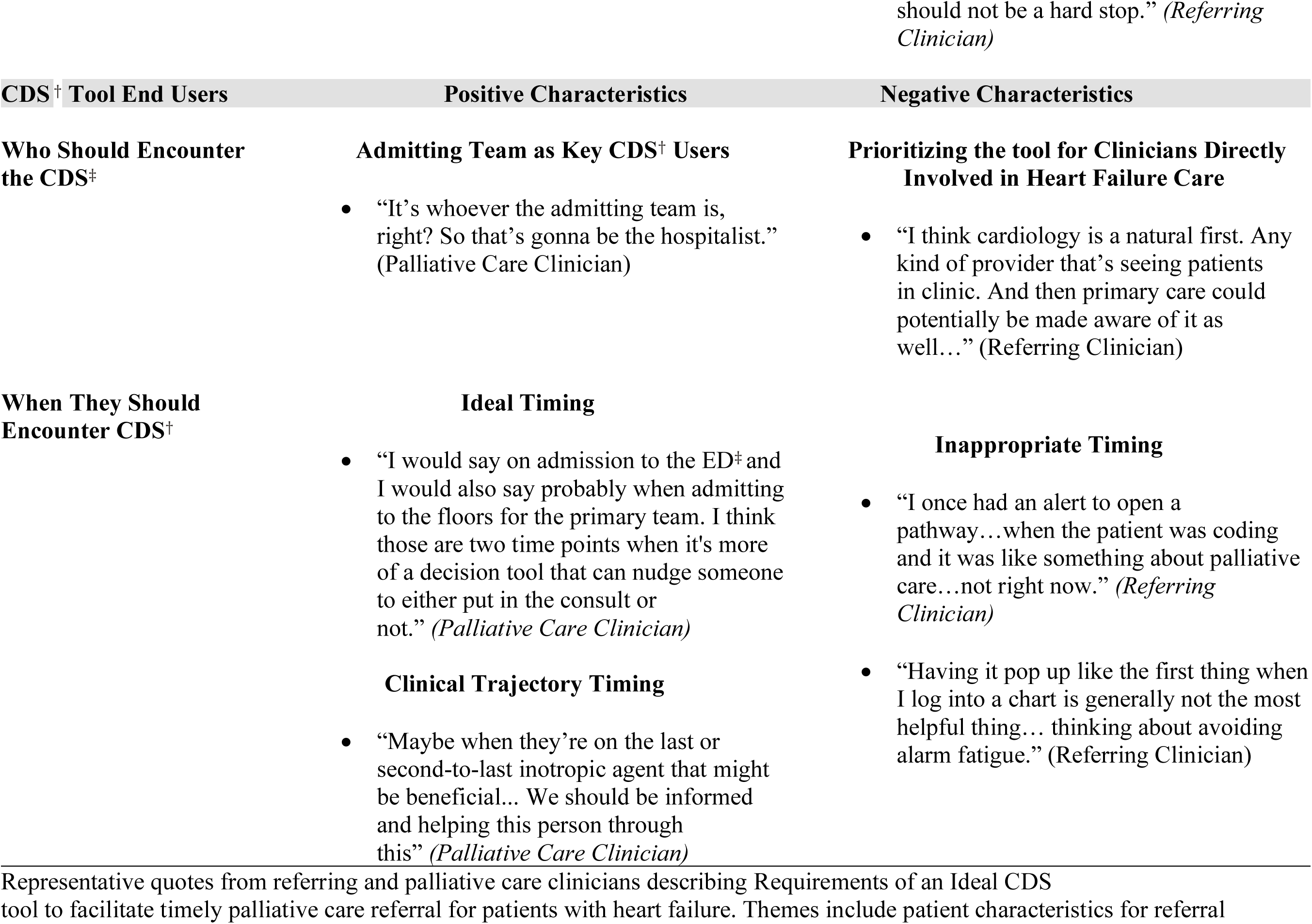
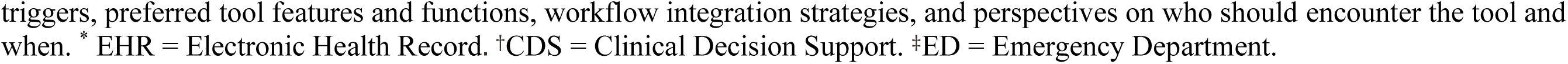
Requirements for an Ideal CDS Tool to Facilitate Palliative Care Referral in Heart Failure.

Clinicians emphasized that referral decisions should always be personalized, reflecting the unique clinical circumstances of each patient (Table 3). They highlighted the need for CDS guidance that accounts for differences in patient status and disease trajectory rather than relying on fixed thresholds.

#### Ideal CDS: features, functionality, integration into workflow

Clinicians provided insights on the desired features, functionality, and integration strategies for a CDS tool designed to facilitate timely palliative care for patients with heart failure (Table 3). Their feedback highlighted priorities in predictive capability, interface design, workflow integration, and support for the palliative care team. Clinicians favored the idea of incorporating predictive risk modeling to proactively identify patients at high risk for palliative needs. Several clinicians recommended the use of a score-based system, while favoring a visual representation of the risk score within the EHR, such as a color-coded system clearly indicating patient risk status. Several also emphasized the importance of flagging repeat admissions for heart failure, which were viewed as an important marker of declining clinical trajectory and heightened risk that patients may be approaching the end of life.

> *“I think that will be really useful… like a number that shows you the mortality… if the number is high, that would definitely make me more inclined to put it as a recommendation in my note.” (Referring Clinician)*

Clinicians stressed that the CDS tool should be integrated into existing workflows, ideally linking with current pathways and order sets while avoiding interruptive alerts, when feasible. They expressed a strong preference for minimizing workflow disruptions, cognitive load, and documentation burden. Suggestions included incorporating pre-filled orders and avoiding additional steps in routine processes and hard stops. Clinicians generally disliked automatic or “trigger” consults, noting that such features could undermine clinician autonomy and potentially overwhelm the palliative care team. Instead, they favored subtle “nudges” and calibrated them to clinical acuity for patients on an individual level.

Clinicians suggested using non-interruptive visual cues within the patient chart as a preferred method of prompting palliative care referrals instead of intrusive pop-ups. One recommended strategy was adding palliative care referral status (e.g., “consulted” or “not consulted”) directly to the prominent left-hand sidebar alongside key patient data, ensuring visibility during routine chart navigation. Others supported the use of color-coded flags, to subtly indicate palliative care eligibility based on specific clinical criteria, such as reduced ejection fraction, multiple hospital admissions, or repeat emergency department visits.

> *“In my ideal scenario, this patient would be flagged because they’ve been readmitted four times for the same condition. And then I could click into it, and then I could do all the things.” (Referring Clinician)*

In addition to supporting patient identification, clinicians highlighted opportunities to streamline the palliative care team’s workflow through the CDS tool. Suggested features included information prompts for referring providers to address often overlooked questions, such as whether the palliative care team has been introduced to the patient and family, and whether there are any pending tests relevant to palliative care planning. Clinicians also recommended that consult orders should auto-populate with critical clinical and contextual information, including code status, prior hospitalizations, and whether goals-of-care conversations had previously occurred.

Clinicians desired a referral pathway from inpatient to outpatient palliative care to ensure greater continuity of care and longitudinal follow-up (Table 3). They noted that outpatient referral could offer patients and families more time to process information, reduce emotional distress, and facilitate greater family involvement. Outpatient settings were also viewed as a strategy to alleviate the burden on the inpatient palliative care team with limited capacity.

#### Who should encounter the tool and when

Clinicians identified a broad range of potential end users for the proposed CDS tool, including cardiology teams, the cardiac ICU, and primary inpatient teams responsible for placing orders during hospitalization, as well as admission and ED teams. Specialists such as nephrologists were also noted, given their role in managing heart failure. In addition to physicians, clinicians emphasized including nurses, case managers, and social workers. These non-physician clinicians were viewed as essential for their sustained interactions with patients, insights into patient needs, and awareness of social context and care coordination, positioning them to recognize non-clinical factors relevant to palliative care.

> *“The nurse knows the family the best, the nurse sees the patient the most, they know the patient the best.” (Referring Clinician)*

Clinicians also emphasized that the timing of the CDS tool’s display is important to its usability and acceptance (Table 3). They agreed that prompts should appear at natural clinical decision points, such as during ED or hospital admission, when clinicians are already reviewing information and making care plans. Several also noted that the tool could reappear at major clinical transitions, including ICU transfers or consideration of advanced therapies like inotropes or LVAD, when prognosis discussions are most relevant. Clinicians preferred that the tool fire within routine workflow steps (i.e., during order entry or documentation) rather than as an intrusive pop-up upon chart login.

> *“I would say like on admission to the ED and when admitting to the floors for the primary team. I think those are two kind of time points when it’s more of a decision tool that can nudge someone to either put in the consult or not.” (Referring Clinician)*

## DISCUSSION

Using the UFIT framework, this study identified critical needs, contextual factors, and requirements for designing a CDS tool to promote timely palliative care for people with heart failure. By engaging key stakeholder clinicians, we sought to understand how patients with heart failure are engaged with palliative care and where a CDS tool could best support earlier referrals (Figure 1). Our findings provide actionable insights into current processes, perceived barriers and facilitators to CDS use, and user preferences for CDS design and integration into existing workflow.

Critical needs addressed knowledge gaps and perceived barriers to timely palliative care. Persistent barriers included uncertainty about when to refer, driven by both prognostic uncertainty and limited clinician knowledge regarding the distinction between palliative care and hospice, as well as the broader scope of palliative services. These findings align with previous studies that have documented similar challenges, including prognostic uncertainty and limited awareness or knowledge of palliative care, as common barriers to timely palliative care.^23–25^

Our findings highlight that identifying the optimal functionality, timing, and placement of a CDS tool within clinical workflows is essential to promoting timely palliative care. Although default “trigger” consults and interruptive tools can be salient and have been shown in some cases to increase palliative referral rates,^26, 27^ studies in the broader CDS literature consistently demonstrate that untailored or poorly timed tools disrupt workflow, contribute to alert fatigue, and rarely improve patient outcomes.^28–30^ Moreover, such designs may be perceived as coercive, administratively driven, and threatening to clinician autonomy.^31^ Prior work has also shown that inappropriate placement of CDS tools is a major barrier to clinician adoption, as tools triggered outside natural workflow steps are often perceived as disruptive or burdensome.^23, 24^ There are also risks associated with these design features. Both our participants and previous research note that limited staffing in palliative care is a persistent problem, and teams may lack the capacity to handle the increased volume of consults that could occur if tools use automatic default “trigger” consults or untailored interruptive designs.^32^

Interruptive alerts need not be abandoned altogether. When used selectively and paired with non-interruptive, workflow-integrated CDS components, they can enhance visibility for high-acuity cases while minimizing overall alert burden. To improve both usability and clinical impact, a tiered recommendation approach could be employed, whereby the level of workflow interruption corresponds to clinical acuity.^33^ Although clinicians in this study did not explicitly describe a tiered approach, their emphasis on tailoring recommendations to patient context aligns with the concept of tiered CDS recommendations, in which guidance is calibrated to clinical acuity. This approach allows high-priority patients to prompt more prominent alerts or recommendations, while lower-risk cases receive more subtle cues. Such stratification reduces unnecessary alerts, minimizes information overload, and prevents alert fatigue by ensuring that clinicians only encounter clinically relevant and actionable notifications.^33^ With careful end-user engagement and thoughtful design, interruptive alerts can be tailored to remain clinically impactful while reducing burden and fostering provider acceptance.^35–37^

This study has several limitations. First, data were collected from clinicians at a single academic health system, and we specifically focused on the inpatient setting, which may limit generalizability to other practice environments with different workflows or referral practices. Second, although we aimed to sample a diverse range of clinicians, our participants may represent individuals with greater interest or experience in palliative care than the broader provider population. Third, because all interviews were conducted within one health system using a single EHR, our findings reflect the alerting capabilities, pop-up functions, and workflow constraints of that specific platform; other EHR systems may offer different or more efficient mechanisms for notifying clinicians, which could influence perceptions of CDS feasibility or design preferences. Lastly, while we captured rich insights into workflows, preferences, and perceived needs, the actual usability and effectiveness of the CDS tool will require further testing in real-world clinical settings.

## Conclusion

We applied the UFIT framework to identify the needs, contextual factors, and design requirements for a CDS tool aimed at improving timely palliative care for patients with heart failure. Our findings emphasize the importance of embedding CDS into existing clinical workflows, minimizing disruption, and incorporating stakeholder input early in the design process. These results will directly inform the development and pilot testing of the CDS tool, to enhance palliative care delivery and support more consistent, timely palliative care among heart failure populations.

## Data Availability

The data that support the findings of this study are not publicly available due to ethical and privacy restrictions involving human subjects. De-identified data may be available from the corresponding author upon reasonable request.

## ACKNOWLEDGEMENTS

None.

